# Genetic Variation at 19q13.33 confers colorectal cancer risk through the interaction of mucosal expression of FUT2 and plasma vitamin B12 levels

**DOI:** 10.64898/2026.07.10.26357741

**Authors:** M Allan, V Rajasekaran, X. Li, B.T Harris, K Donnelly, M Walker, E. Miedzybrodzka, H. Wang, K Myant, F. V Din, S.M Farrington, M. G. Dunlop

## Abstract

**Introduction:** Genome-wide association studies have identified a common variant at chr19q13.33 within the *FUT2* locus as a determinant of colorectal cancer (CRC) susceptibility, with each risk allele conferring an approximately 7% increase in risk (OR 1.07, P = 6.11×10⁻¹⁰). This locus regulates expression of FUT2, a fucosyltransferase involved in α-1,2 glycosylation, and has also been associated with circulating vitamin B12 (B12) concentrations in genome-wide studies.

**Objective:** To determine whether FUT2 influences CRC-risk through effects on circulating B12 and to test causal relationships across genetic, experimental, and clinical data.

**Methods:** We performed summary-data-based Mendelian randomisation using FUT2 eQTLs from GTEx colon tissue and genome-wide association data for plasma B12 (Generation Scotland), with mediation analysis estimating the proportion of effect mediated by B12. Causal inference was then tested in vivo using *Fut2* knockout and wild-type mice exposed to azoxymethane/dextran sodium sulphate (AOM/DSS), with or without B12 supplementation.

**Results:** Genetically predicted higher *FUT2* expression was associated with lower B12 levels (β = −0.735, SE = 0.110, P = 2.63⁻¹¹) and reduced CRC-risk (β = −0.256, SE = 0.058, P = 5.85 x10⁻⁵). Mediation analysis suggested ∼80% of the effect of FUT2 on CRC risk is mediated via B12. In mice, neither Fut2 deficiency nor B12 supplementation alone induced tumours, but both significantly increased tumour burden under chemical carcinogenic challenge, with comparable effect sizes.

**Conclusions:** B12 is a key mediator of FUT2-associated CRC risk, supported by convergent genetic, experimental, and clinical evidence implicating altered B12 metabolism in colorectal neoplasia susceptibility.

## Introduction

35% of Colorectal cancer (CRC) is characterized by heritable risk, of which approximately 5% is accounted for by highly penetrant inherited mutations, such as germline mutations of *APC*.^1^ The remaining susceptibility is accounted for by numerous common mutations of low penetrance.^2^

Genome wide association studies (GWAS) have identified over 200 independent genetic variants as being responsible for CRC risk.^3^ One of these CRC-risk loci, rs12979278, is a single nucleotide polymorphism (SNP) at 19q13.33 that is associated with a 7% per allele excess risk of CRC (OR 1.07, p=6.11×10^-10^) and maps close to the gene, *FUT2* (fucosyltransferase 2).^4^ Subsequent analysis of expression quantitative trait data identified associations between rs12979278 and *FUT2* expression and summary-data-based Mendelian Randomization (sMR) analysis provided support that SNPs at 19q13.33 influence CRC-risk through differential expression of *FUT2* - lower expression being associated with increased risk.

*FUT2* encodes the enzyme α(1,2)-fucosyltransferase, which is preferentially expressed in epithelial cells within the intestinal mucosa and is responsible for producing the H-antigen in non-blood bodily fluids. The H-antigen serves as a precursor of the ABO blood group antigens, and individuals with these soluble antigens are termed secretors.^5^ As *FUT2* is dominant, one functional allele (Se) confers the secretor phenotype, whereas homozygosity for a non-functional allele (se) result in non-secretor status. Multiple allelic variants underlie these phenotypes, with rs601338 and rs602662 being the most common in Europeans and showing high linkage disequilibrium (95%).^6,7^

Multiple GWAS have also shown *FUT2* genotype to influence vitamin B12 (B12) homeostasis, with non-secretor (se) variants consistently associated with elevated circulating B12 across diverse populations ^8,9,10,11^. B12, obtained primarily from animal foods, requires binding to intrinsic factor, secreted by gastric parietal cells, for absorption in the terminal ileum. While diet and gastrointestinal pathology affect B12 status, genetic factors account for 56–59%, with *FUT2* among the strongest loci, alongside TCN1 and TCN2.^11,12^

An initial GWAS in European females identified rs492602, in strong LD with rs602662 and rs601338, as strongly associated with higher B12 levels (β=–0.09, p=5.36 × 10⁻¹⁷)^5^ a finding subsequently replicated across European, North American, and Asian cohorts.^8,9,10,13^ Observational and MR studies now indicate a positive association between genetically predicted B12 levels and CRC risk, raising the possibility that *FUT2*-mediated alterations in B12 metabolism contribute to CRC development. Clinically, higher serum B12 has been associated with CRC in the B-PROOF trial (OR per SD=1.21, p=0.016)^14^ and MR analyses further support causality. Genetically predicted B12 concentrations were positively associated with digestive system cancers overall (OR=1.12, p=0.003) and CRC specifically (OR=1.16, p=0.001) in UK Biobank and FinnGen,^15^ with additional supportive evidence from a European study.^16^ Together, these findings suggest *FUT2* variants influence B12 metabolism in ways that may contribute to CRC development.

The mechanisms linking *FUT2*, B12, and CRC remain incompletely understood. One proposed hypothesis is through modulation of the gut microbiome. *FUT2* encodes a fucosyltransferase enzyme that regulates α-1,2 glycosylation.^17^ These fucosylated glycans act as attachment sites and nutrient sources for certain bacteria, including *Helicobacter pylori,* thus *FUT2* polymorphisms may influence intestinal microbiome.^18, 19^ The presence of *H. Pylori* in the intestine has been reported to interfere with the release of intrinsic factor, which is needed for B12 absorption.^20^ Experimental data from *FUT2*-null mice and human studies demonstrate intestinal microbial shifts with altered fucosylation, although results are inconsistent.^21–24^

To summarise, GWAS have implicated common variance in *FUT2* as a causal factor in CRC-risk, with reduced expression of *FUT2* in the colorectal epithelium associated with increased risk. Lower *FUT2* expression has been linked to elevated plasma B12 levels, and whilst higher B12 has been associated with increased CRC-risk, the evidence remains inconclusive. *FUT2* genotype has also been correlated with alterations in the composition of the intestinal microbiome in both human and mouse studies, with these changes tending toward a more pathogenic microbial profile. Finally, although current evidence supports an association between the intestinal microbiome and CRC-risk, a definitive causal relationship has yet to be established. Findings to date suggest a complex interplay between *FUT2* genetics, B12 metabolism, and CRC-risk, but direct evidence remains limited. In this study we interrogate this relationship using genetically engineered mouse models and sMR analysis.

## Material and Methods

### Mouse models

All animal experiments were performed at the University of Edinburgh’s Biomedical and Veterinary Sciences facility under UK Home Office licences P02F16F82 and PP8297251, following approval by the University’s Animal Welfare and Ethical Review Board and in compliance with UK legislation. Mice were housed in individually ventilated cages (1–4 per cage) at 21–22 °C with a 12 h light–dark cycle and given ad libitum access to irradiated RM3 food (Special Diets Services, UK) and bottled water.

### *Fut2* mouse models

Cryopreserved B6.129X1-*Fut2^tm1SDo^/*J embryos (Jackson Laboratory) were implanted into C57BL/6J recipients to generate *Fut2^-/-^* mice on a mixed 129X1/SvJ × C57BL/6J background, alongside Fut2^+/+^ controls. In this line, a LacZ cassette replaces the exon encoding the catalytic domain of *Fut2*.^25^

### Fut2 Apc^min/+^ mouse models

*Fut2^-/-^*mice were subsequently crossed with *C57BL/6J-Apc^min/+^* mice from the Sansom Laboratory (CRUK Scotland Institute) to generate the *Fut2 Apc^min/+^*line.

### *Apc ^flox/+^* mouse models

*VilCre^+^Apc^flox/+^* mice from the Myant laboratrory (University of Edinburgh), in which loxP sites flank exon 14 of the *Apc* gene, were used for conditional *Apc* recombination experiments. Recombination was induced by tamoxifen administration via oral gavage at a dose of 100mg/kg for five consecutive days. Control animals consisted of *Apc^flox/+^* littermates lacking Cre recombinase or *VilCre^+^Apc^flox/+^* mice that did not receive tamoxifen treatment.

### Genotyping of mice

Ear clips from 21-day-old Fut2 mice were lysed in Viagen Direct PCR reagent with proteinase K (20 µg mL⁻¹), incubated at 55 °C for 14 h and 85 °C for 1 h, centrifuged (10,000 *g*, 30 s), and stored at –20 °C. PCR was performed using 1 µL lysate with primers from Integrated DNA Technologies (Supplementary Tables 1–3) following Jackson Laboratory protocols. *Fut2* genotypes were resolved on 3% agarose gels.

Genotyping of the *Fut2 Apc*^min/+^ mice and the *Apc^flox/+^* mice was carried out by Transnetyx.

### Experimental design

*Fut2^-/-^* and *Fut2^+/+^* mice were assigned to one of six experimental groups. A control group (A) which was aged to 22 weeks, received normal chow and water and no further intervention. A chemical carcinogen group (B) which received a single intra-peritoneal injection of 10mg/kg AOM at 12 weeks of age followed by a 1 week course of 1% DSS in drinking water at 13 weeks. A B12 supplementation group (C) which received 100mg/kg/day of B12 in drinking water from age 10 to 22 weeks. A B12 chemical carcinogen group (D) which received both chemical carcinogens and B12 supplementation in drinking water, as described above. A fifth cohort were treated with 100mg/kg/day of B12 in the drinking water from weaning (week 6) until 1 year at which point they were culled (E). An additional ageing cohort received normal chow and water, no intervention and were culled at 2 years (F). All groups had both male and female mice from each genotype (Fig 1).

**Figure 1.**
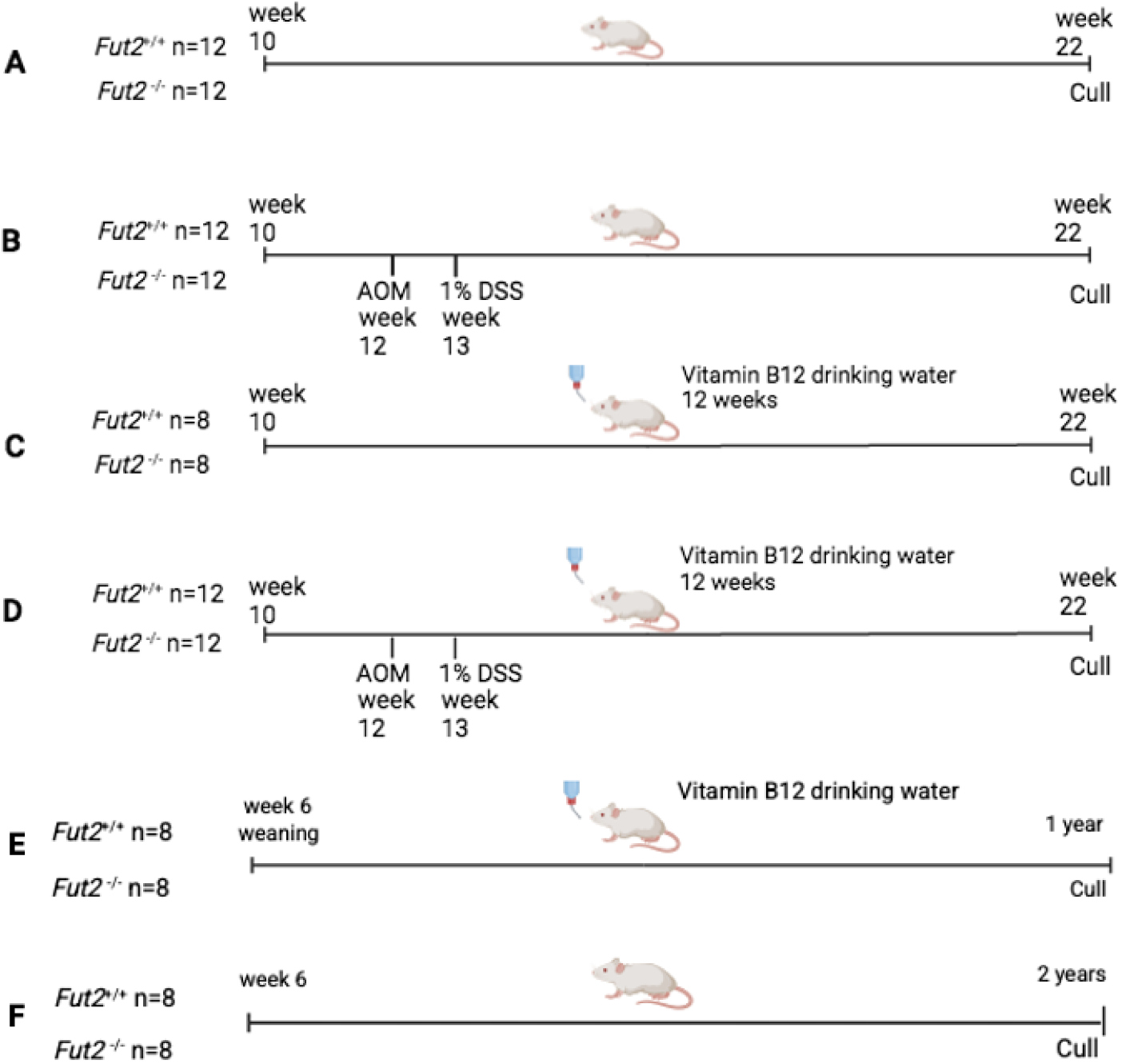
Schematic representation of the four different cohorts of mice experiments. A control group (n=24), B a chemically induced CRC group, AOM (10mg/kg) intraperitoneal injection and 1% DSS in drinking water for 7 days (n=24) C B12 supplementation group (100mg/kg/day) for 12 weeks (n=16) D B12 supplementation and chemically induced CRC group (n=24). E B12 supplementation group (100mg/kg/day) for 1 year. F ageing group given normal chow and water and aged to 2 years.

Mice were monitored twice daily, including daily weights during the 1 week course of DSS administration and for up to 5 days afterwards. Should any mice have shown a change in health during this time frame they were culled and replaced. This included >20% weight loss over 48 hrs, significant rectal bleeding, rectal prolapse or motionless with hunched posture (parameters described in Supplementary Table 4). Additionally, mice were given a health score just prior to culling using the same parameters. Power calculations were performed to determine the required sample size necessary to detect a significant difference based on *Fut2* expression (n=8). Additional mice (n=4) were assigned to chemical carcinogen experimental groups due to the unknown effects of the AOM/DSS concentrations and possible early death associated with chemical inflammation.

*Fut2 Apc ^min/+^*mice received normal chow and water and aged until they showed sign of the above clinical parameters mentioned (Supplementary Table 4), no intervention was undertaken on these mice.

*Apc^flox/+^*Cre^+^ mice treated with tamoxifen were divided into two groups (n=10 per group) and received either B12 supplementation in drinking water from weaning (approximately weeks 5–6) or standard drinking water. These mice were culled at week 22, or earlier if they exhibited predefined ill-health criteria (see Supplementary Table 4). Control groups, also treated with B12 from weaning, consisted of *Apc^flox/+^*Cre^−^ mice administered tamoxifen (n=10) and *Apc^flox/+^*Cre^+^ mice not treated with tamoxifen (n=10).

In all experimental cohorts and mouse lines both male and female mice were used.

### Azoxymethane /Dextran Sulphate Sodium

To induce inflammation-associated tumorigenesis, mice received a single intraperitoneal injection of azoxymethane (AOM, 10 mg/kg body weight) at 12 weeks of age, followed by a one-week course of 1% dextran sodium sulphate (DSS) in the drinking water at 13 weeks of age. DSS was prepared in sterile water at a 1% concentration.

### B12 preparation and administration

B12 was purchased in powder form (MilliporeSigma, Darmstadt, Germany) and was administered in the drinking water of experimental mice from 10 weeks of age in *Fut2* mice and from 6 weeks of age in the *Apc^flox/+^*mice until study end. Bottles were prepared with distilled water containing B12 at 100 mg/kg body weight/day, based on an estimated daily water intake of 5 mL per mouse. Drinking water was replaced weekly, and dark-coloured bottles were used to protect the light-sensitive B12.

### Mouse Tissue Collection and preparation

Mice were culled by cervical dislocation, and intestines were dissected from pylorus to terminal ileum (small intestine) and ascending colon to anus (colon). Intestinal segments were mounted on wooden skewers, fixed in 10% NBF (24 h, RT or 72 h, 4°C), and stored in 70% ethanol.

### Methylene Blue Staining

Whole intestines were dipped five times in 0.2% methylene blue and de-stained in 70% ethanol at 4°C for 7–10 days (replacing ethanol every 2–3 days). Brightfield images were acquired using a Pippin Mesoscope (0.5×), and polyps identified by intense blue staining were measured with LASX software.

### Swiss Rolling, embedding and sectioning

Intestinal samples were prepared using a “Swiss-roll” technique to provide cross-sections that incorporate the entire length of each sample.^26^ Intestines were opened longitudinally, laid flat (epithelium up), and rolled along their length (Swiss-roll). Rolls were paraffin-embedded, sectioned onto Superfrost slides, and subsequently used for H&E and immunostaining.

### Histology and Immunohistochemistry (IHC)

Paraffin-embedded tissues were deparaffinised, rehydrated, and peroxidase-blocked. Antigen retrieval was performed using either 0.1% trypsin in PBS (FUT2 lectin) or heat-induced citrate buffer (IHC) in a pressure cooker (10–15 min), followed by PBS-T washes. Slides were permeabilised (0.5% Triton X-100, 20 min), blocked (1% BSA + 5% goat serum, 1 h), and incubated with primary antibodies or biotinylated UEA-I lectin (Supplementary Table 5) at 4°C or 37°C as appropriate, followed by secondary antibodies. 3,3′-diaminobenzidine (DAB) was used for signal development, nuclei counterstained with Harris’ haematoxylin, and slides were dehydrated and mounted.

### β-Galactosidase Staining

Intestinal tissues from *Fut2⁻/⁻* mice were fixed on ice for 2 h in 10% NBF with glutaraldehyde and NP-40, washed in PBS, and incubated in X-gal staining solution in the dark for 24 h (Supplementary Table 6). Tissues were post-fixed in 10% NBF overnight, Swiss-rolled, sectioned at 10 µm, and counterstained with Neutral Red.

### Imaging and Analysis

H&E and IHC slides were scanned with a Nanozoomer Hamamatsu slide scanner and analyzed using NDP Viewer software (NDP .view2). Images were interpreted by a single blinded investigator, with abnormalities reviewed by two additional researchers. Additionally, selected H&E slides were cross-validated by a specialist intestinal pathologist.

For analysis of IHC, quantification of positively stained cells was performed using QuPath (v0.5.1).^27^ As illustrated in Fig.2, representative regions were first selected to define stain vectors for DAB and Harris haematoxylin in each image. Regions of interest were then delineated using the markup tool, and positive cell detection was carried out based on the cell DAB optical density (OD) mean parameter. QuPath reported both the total number of cells and the number of positively stained cells within the annotated regions (Fig. 2).

**Figure 2.**
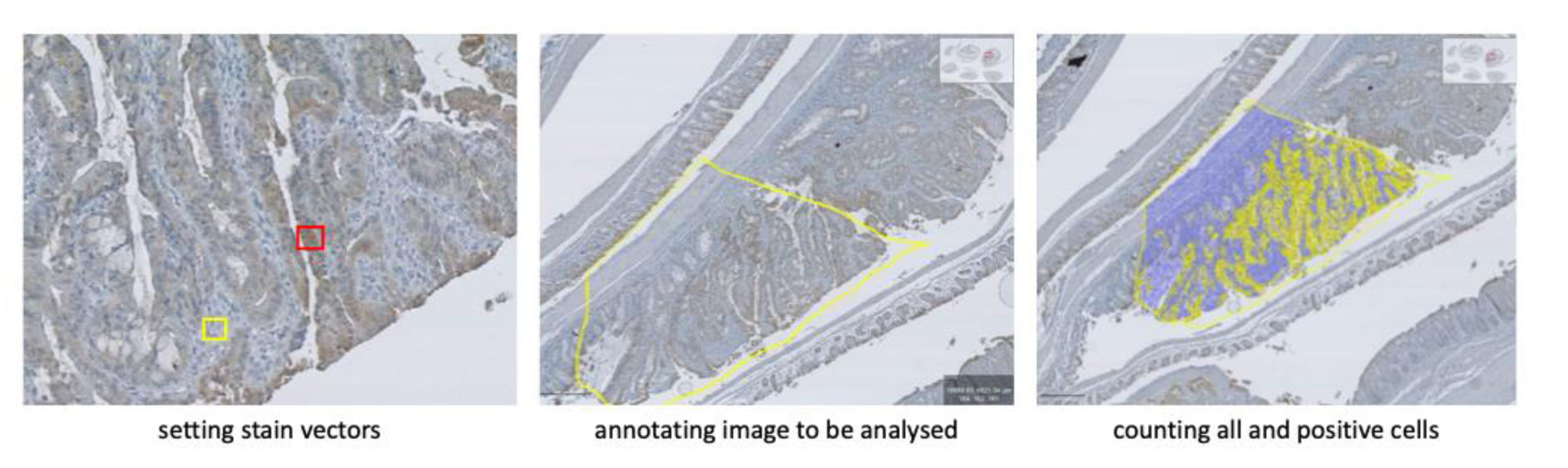
Q-path quantitative analysis of cells stained positive for E-cadherin with DAB. Stain vectors for haematoxylin (yellow box) and DAB (red box) are set for each image. Area to be analysed defined with markup tool. Positive cell detection tool set for analysing “cell DAB OD

All statistical analyses and data visualizations were performed using GraphPad Prism (v10.4). Unless otherwise specified, *p-*values were calculated using an unpaired two-tailed *t*-test with Welch’s correction. Bar graphs are presented as mean ± standard error of the mean (s.e.m.), unless stated otherwise.

### Summary-based Mendelian Randomisation (SMR) and Mediation Analysis Study population and datasets

Observational associations between *FUT2*, B12, and colorectal cancer (CRC) were evaluated in the UK Biobank (UKB), a large prospective cohort with extensive genotypic and phenotypic data.^28^ CRC cases were ascertained through national cancer and death registries and Hospital Episode Statistics, defined as malignant neoplasms of the colon, rectum, or rectosigmoid junction (ICD-9/ICD-10). After excluding participants with missing genotype data, the analysis included 9,276 cases and 477,069 controls.

Causal relationships were assessed using SMR. The largest CRC GWAS dataset among European populations to date was used, including 78,473 cases and 107,143 controls of European ancestry.^4^ This analysis integrated expression quantitative trait loci for *FUT2* in transverse colon mucosa (GTEx v8), genetically predicted serum B12 (from a Generation Scotland GWAS), and CRC-risk summary statistics from the largest European-ancestry CRC GWAS to date (78,473 cases, 107,143 controls; Colon Cancer Genetics Group).^3^

### FUT2-B12-CRC association and interaction analysis

Polygenic risk scores were conducted for *FUT2* (or B12) based on SNPs (*p*<5E-8, r^2^<0.1) by summing the product of the number of effect alleles (0, 1 and 2) and corresponding effect size on *FUT2* expression in colon tissue (or B12 level) genetic liability for each effect variant to indicate the genetically predicted *FUT2* expression (or B12 level) in UKBB. Then, the independent effects of *FUT2* gene types, genetically predicted *FUT2* expression, and genetically predicted B12 level (continuous and categorical) on CRC risk were estimated using Logistic regression. Next, the interactions of B12 level (continuous) with *FUT2* gene types on CRC risk were estimated. Age, sex, and the first 10 genetic principal components were adjusted in the model.

### Summary-data-based Mendelian randomization analysis

For further evaluating the causal associations between *FUT2*, B12, and CRC risk, the SMR analyses were conducted.^29^ The heterogeneity in dependent instruments (HEIDI) test was employed to distinguish exposure (*FUT2* or B12) that were associated with the risk of CRC owing to a shared variant rather than genetic linkage. The SMR software (SMR v1.3.1) was used to perform SMR and HEIDI test.^29^ A *p-*value <0.05 was defined as the statistical significance level for SMR. The *p-*value of HEIDI test >0.05 indicated that the association of exposure and CRC was not driven by linkage disequilibrium.

### Mediation analysis

Linear regression was used to assess the association between genetically predicted *FUT2* expression and B12 levels, while logistic regression evaluated their associations with CRC risk. Mediation analysis was conducted using the “mediation” R package^30^ to estimate the average causal mediation effect (ACME), average direct effect (ADE), and proportion mediated.

A two-step SMR approach was applied to assess mediation. First, genetic instruments for *FUT2* expression were used to estimate its causal effect on B12. Second, instruments for B12 were used to estimate its causal effect on CRC risk. The indirect effect of *FUT2* on CRC via B12 was calculated using the product-of-coefficients method,^31^ with standard errors estimated by the delta method via the “RMediation” R package.^32^

## RESULTS

### Mediation MR analysis indicates that a significant proportion of *FUT2’s* effect of CRC-risk is mediated by B12

SMR and mediation analyses were conducted to interrogate the role of B12 in mediating *FUT2-*associated CRC risk. SMR enables assessment of the predicted direct causal effect of B12 on CRC risk in humans while minimizing bias from confounding factors.

The primary analysis used a CRC case–control dataset from UK Biobank, and findings were replicated in the largest European CRC GWAS dataset, integrating *FUT2* expression data from GTEx with serum B12 GWAS summary statistics from Generation Scotland.

The genetically predicted expression level of *FUT2* in colon tissue was inversely associated with CRC risk (β=-0.039, *p*=0.041), whereas the genetically predicted B12 level was positively associated CRC risk (β=0.094, *p*=0.007) (Table 1). A marginal interaction effect between *FUT2* expression and B12 level on CRC risk was found (*p*_interaction_=0.04) (Table 1), but no significant association between *FUT2* (or B12) and CRC risk was found after stratifying B12 levels by quartiles (or *FUT2* expression) (*p*>0.05)(Table 2).

**Table 1.** The associations and interaction of FUT2 gene types and vitamin B12 level on CRC-risk in UK Biobank. Genetically predicted FUT2 expression is inversely associated with CRC-risk (β=-0.039, *p*=0.041). Genetically predicted vitamin B12 level is positively associated the CRC-risk (β=0.094, *P*=0.007). Marginal interaction effect between *FUT2* expression and vitamin B12 level on CRC-risk (*p*interaction =0.04)

| Main effect |  |  |  |  |  | Interaction effect |  |
| --- | --- | --- | --- | --- | --- | --- | --- |
| *FUT2* |  |  | Vitamin B12 |  |  | *FUT2* * B12 (continuous) |  |
| *FUT2* gene types | Beta | *p* | B12 classifications | Beta | *p* | Beta | *p* interaction |
| rs35866622_C_T<sup>a</sup> | -0.052 | 5.35E-04 | Continuous | 0.094 | 0.007 | -0.024 | 0.616 |
| rs601338_G_A<sup>b</sup> | -0.045 | 2.47E-03 | Dichotomous | 0.042 | 0.048 | -0.012 | 0.798 |
| rs281377_T_C<sup>c</sup> | -0.043 | 4.50E-03 | Tertiles | 0.022 | 0.090 | 0.011 | 0.832 |
| *FUT2* expression | -0.039 | 0.041 | Quartiles | 0.23 | 0.017 | -0.128 | 0.040 |

Adjusted for age, sex and the top 10 genetic principal components.
Vitamin B12, genetically predicted B12 level.
*FUT2* expression, genetically predicted *FUT2* expression in colon tissue.
<sup>a</sup> Effect allele: C
<sup>b</sup> Effect allele: G
<sup>c</sup> Effect allele: T
Note: the effect allele predicted higher level of *FUT2* expression.

**Table 2.**
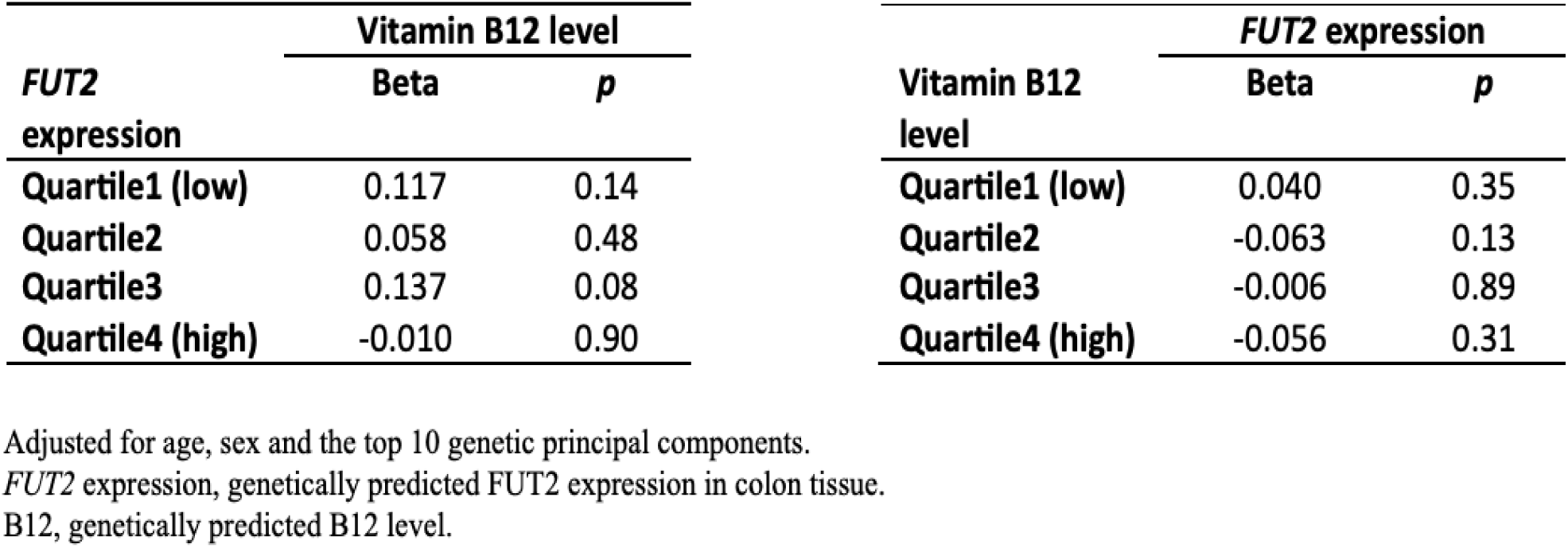
No significant association between *FUT2* (or vitamin B12) and CRC-risk was found after stratifying vitamin B12 levels by quartiles (or *FUT2* expression by quartiles) in UKB. A The association between vitamin B12 (continuous) and CRC-risk stratified by *FUT2* expression (quartiles). B The association between *FUT2* expression (continuous) and CRC-risk stratified by vitamin B12 (quartiles).

SMR analysis of the CRC-GWAS data supported the causal associations of *FUT2* expression with B12 (b;-0.735[SE]=0.110, p=2.63×10-11) and with CRC-risk (b;-0.256 [SE]=0.058, p=5.85 x10-5) and no heterogeneity was found (*p*_HEIDI_>0.05) (Table 3).

**Table 3.**
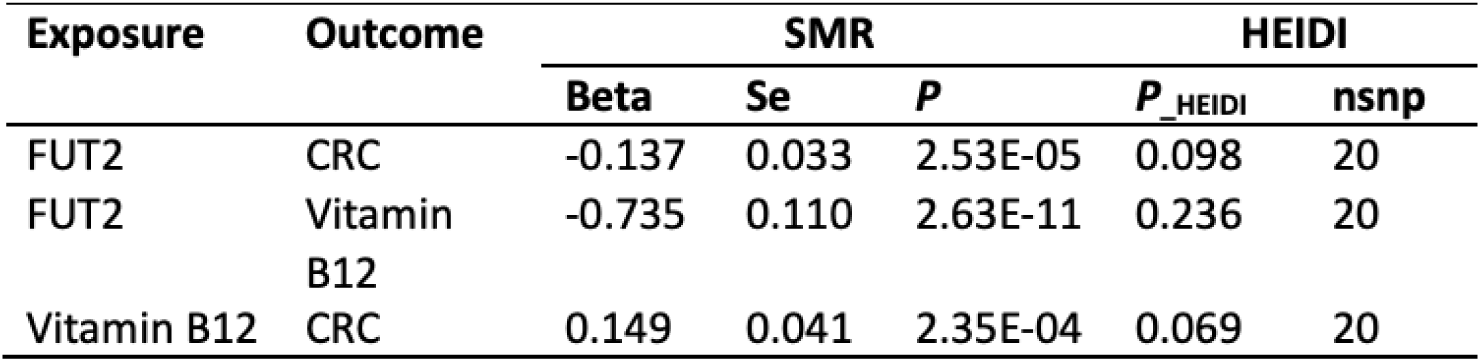
Summary results from SMR study using CRC-GWAS data and HEIDI test. *FUT2* expression was causally associated with vitamin B12 (b;-0.735[SE]=0.110, p=2.63×10-11) and with CRC-risk (b;-0.256[SE]=0.058, p=5.85 x10-5).

Mediation MR analysis using UKB data, showed that genetically predicted B12 levels mediated 31.5% of the association of genetically predicted *FUT2* expression with CRC-risk (ACME p=0.04) (Table 4).

**Table 4.**
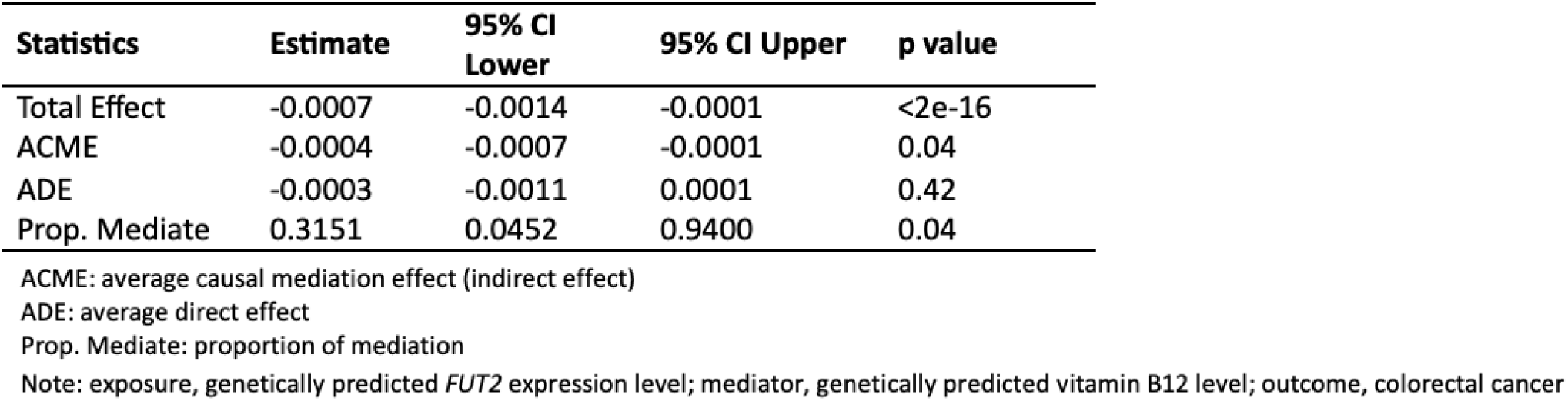
Mediation statistics of Vitamin B12 level on the association between *FUT2* expression and CRC in UK Biobank. . 31.5% of *FUT2* expression on CRC -**risk** is mediated by vitamin B12 (p=0.04).

The mediation effect was further supported by two-step SMR analysis using the CRC-GWAS data, and genetically predicted levels of B12 mediated 80% of the effect of *FUT2* expression on CRC-risk. (Fig. 3).

**Figure 3.**
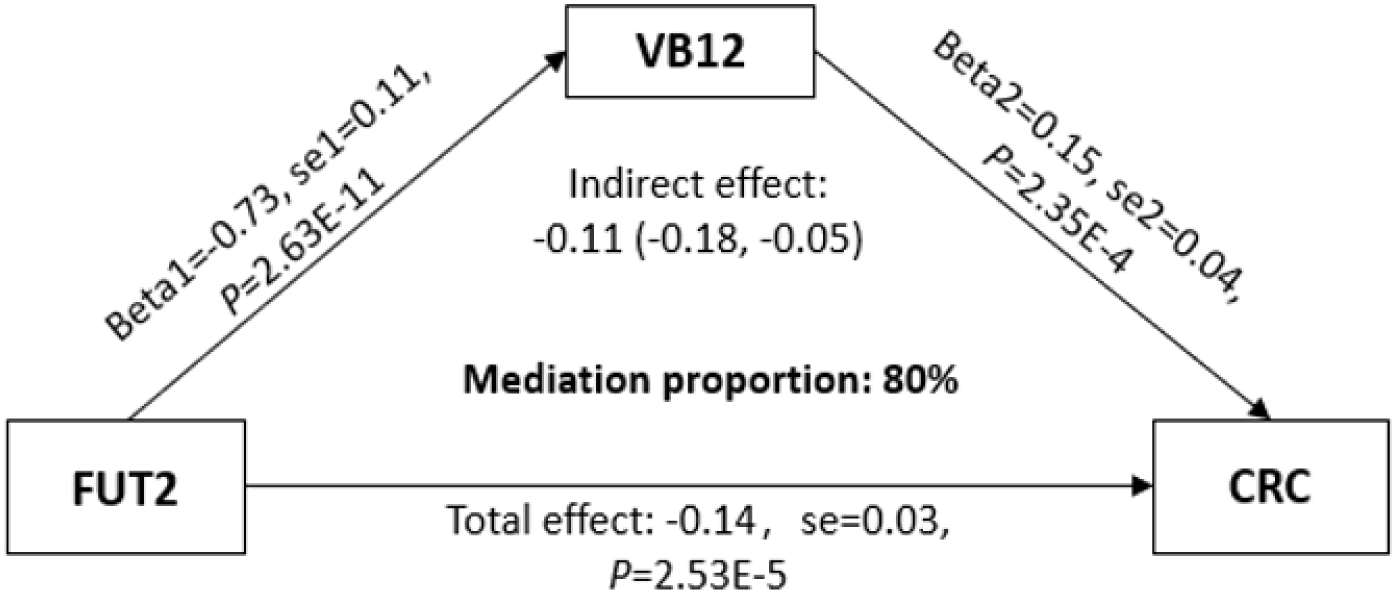
Two-step SMR analysis using CRC-GWAS data. 80% of the effect of *FUT2* expression on CRC-risk is mediated by vitamin B12, indicating that higher B12 increases CRC-risk.

### *Fut2^-/-^* mice exhibit reduced litter sizes and reduced survival at 30-day analysis

To functionally assess the 19q13.33 eQTL CRC risk locus, a *Fut2^-/-^* mouse line was generated. Homozygous deletion of *Fut2* gene was confirmed by PCR and immunohistochemical staining of intestinal tissue. *Fut2^-/-^*mice exhibited β-galactosidase staining in goblet cells of the colon, indicating *LacZ* reporter expression, whilst *Fut2^+/+^* mice stained positively for Lectin Ulex europaeus agglutinin I, which binds α1,2-linked fucose (Fig. 4A).

**Figure 4.**
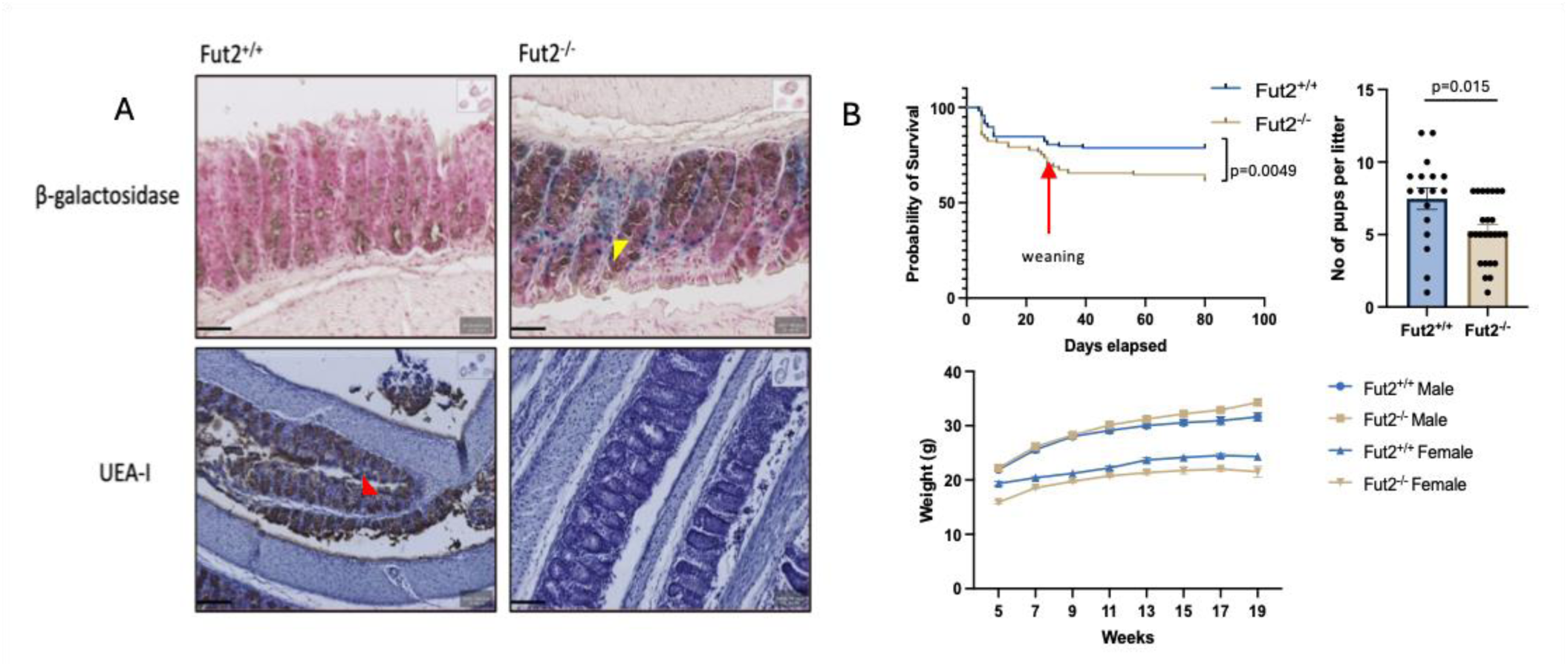
Validation of *Fut2* knockdown and associated phenotypes. A β-galactosidase staining (blue, yellow arrow) indicates the presence of the *LacZ* cassette, while Ulex europaeus agglutinin I (UEA-I) staining (brown, red arrow) marks α-L-fucose and functional Fut2 expression. Scale bars, 50 µm. B Kaplan–Meier survival curve (sexes combined), significant reduction in survival between day 20-40 at time of weaning from dams in *Fut2-/-* mice (red arrow); log-rank (Mantel–Cox) test, *p*=0.0049. Litter size comparison; unpaired *t*-test with Welch’s correction, *p*= 0.015. No significant genotype-dependent differences in body weight within sex; error bars show SEM, *t*-test of area under the curve, p> 0.05.

*Fut2^-/-^* mice were viable and showed no gross physical or behavioural abnormalities consistent with previous literature.^25^ However, whilst most studies have reported *Fut2^-/-^* mice to be fertile and normal in size, this study observed a significant reduction in litter size compared to *Fut2^+/+^*mice (7.47 vs 5.24, p=0.015) (Fig 3B). Survival analysis over the first 80 days of life revealed a significant decrease in survival among *Fut2^-/-^* mice (p=0.0049), particularly between days 20-40, corresponding to weaning and the time of separation from the dam (Fig 4B). No significant difference in bodyweight were observed between genotyped mice until 19 weeks of age (Fig. 4B).

### Loss of *Fut2* increases tumour burden in a CRC inflammatory murine model

Mice in the control group (Group A; *Fut2^-/-^* n=12, *Fut2^+/+^* n=12), maintained on normal chow and water, exhibited no abnormal phenotypes or CRC tumours at 22 weeks as confirmed by methylene blue staining and H&E analysis. This absence of pathology was also observed in all mice aged to 2 years irrespective of genotype (Group F; *Fut2^+/+^* males n=5, *Fut2^+/+^* females n=4, *Fut2^-/-^* males n=4, *Fut2^-/-^* females n=4). In contrast, when treated with AOM/DSS, *Fut2^-/-^*mice (Group B; *Fut2^-/-^* n=12, *Fut2^+/+^*n=12), developed colonic pathology prior to the 22-week endpoint, including rectal bleeding, hunched posture, and, in one case, rectal prolapse, resulting in significantly higher clinical scores compared with *Fut2^+/+^* mice (2.17 vs. 0.25, p=0.008, Fig 5) and reduced survival (p=0.05), with three mice requiring early culling. Methylene blue staining and macroscopic imaging confirmed tumour formation in both genotypes, but *Fut2^-/-^* mice exhibited a higher tumour number per colon (4.92 vs. 2.0, p=0.019), greater total tumour burden, defined as the maximum combined diameter of all tumours in a single colon (11.29 mm vs. 2.0 mm, p=0.0009), and elevated tumour burden scores (6.38 vs. 2.85, p=0.0095), a validated metric derived from the tumour number and maximal tumour diameter.^33^ Tumours were predominantly located in the rectum and mid colon in both genotypes (Fig. 5)

**Figure 5.**
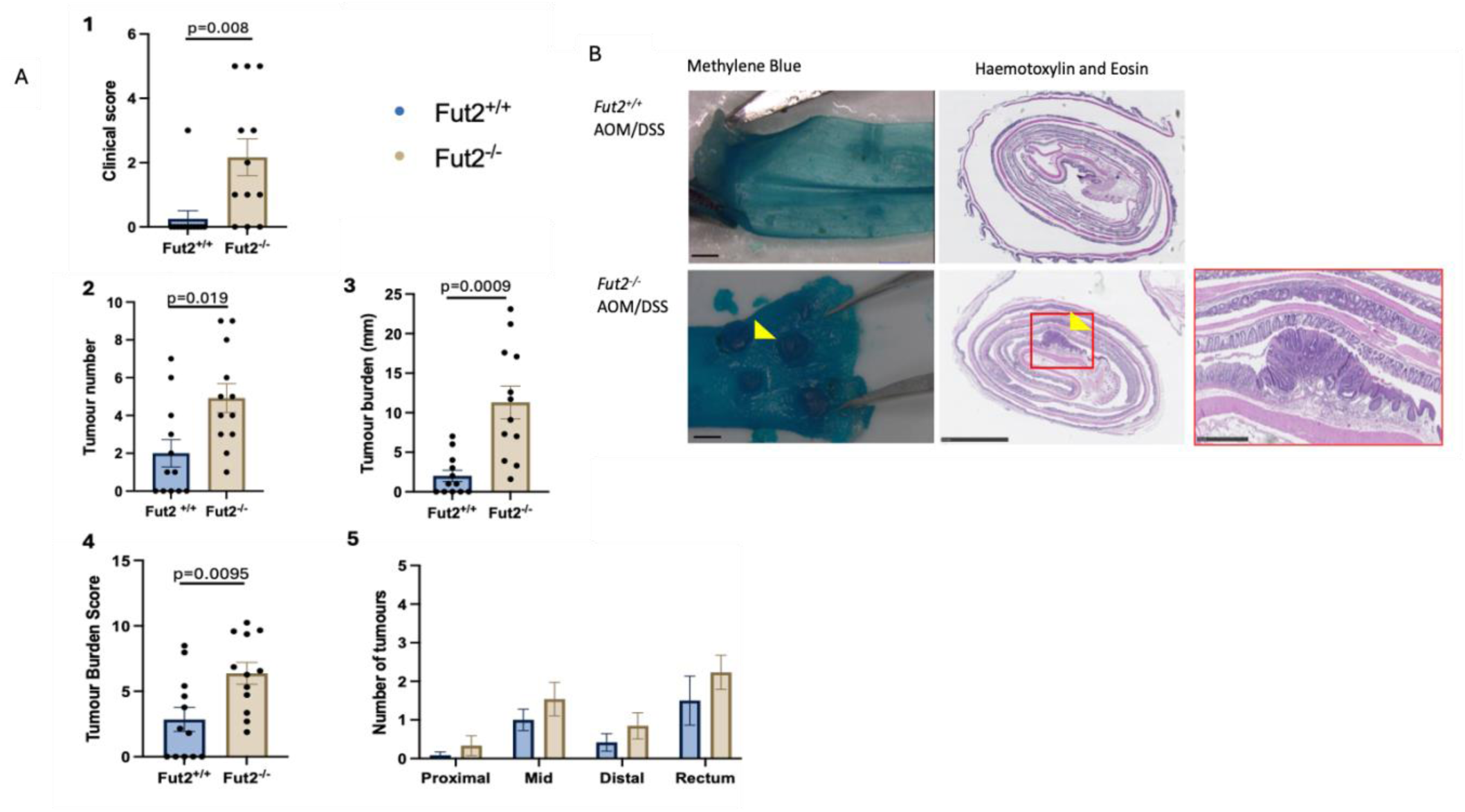
Validating *Fut2* as a CRC-risk locus in an inflammatory CRC murine model. A 1. Clinical score at the time of culling (p=0.008), 2. tumour number (p=0.019), 3. tumour burden (p=0.0009), 4. tumour burden score (p=0.0095) and 5. tumour location with results shown as mean with standard error of mean in the case of bar charts. The tumour burden is the sum of the maximum diameters of all the tumours in a single colon specimen (mm). The tumour burden score calculated as √((Max Diameter)2 +(Total tumour number)2) and therefore is a derivative of tumour burden and number. P-values calculated with unpaired T-test with Welch’s correction for all bar charts and calculated using log rank (mantel-cox) test in survival analysis. B Sections of colons from *Fut2+/+* and *Fut2-/-* mice treated with AOM/DSS, stained with methylene blue to highlight the colonic tumours. Haematoxylin and Eosin (H&E) sections from the same mice to highlight the cross-sectional view. Yellow markers denote examples of tumours. Scale bars set at 2mm with the exception of the red magnified image at 500μm.

Paraffin-embedded intestinal Swiss-rolls were stained for E-cadherin and UEA-I Lectin. E-cadherin, a tumour suppressor critical for adherens junctions, is often lost during tumour progression.^34^ Quantitative analysis using QuPath revealed a significantly greater number of cells stained positive for E-cadherin in *Fut2^+/+^* mice (n=7) compared to *Fut2^-/-^* mice (n=8) treated with AOM/DSS in both tumour ((36.31% (*Fut2 ^+/+^*) vs. 11.16% (*Fut2^-/-^),* p=0.0085) and normal epithelium (33.77% (*Fut2 ^+/+^*) vs. 14.85% (*Fut2^-/-^),* p=0.0062)(Fig. 6). UEA-I Lectin staining, which detects α-1,2-fucosylated structures, demonstrated a significant reduction in *Fut2* expression in tumour epithelium of *Fut2^+/+^*mice compared with adjacent normal tissue (p=0.0008), indicating local loss of fucosylation during tumour development (Fig 7).

**Figure 6.**
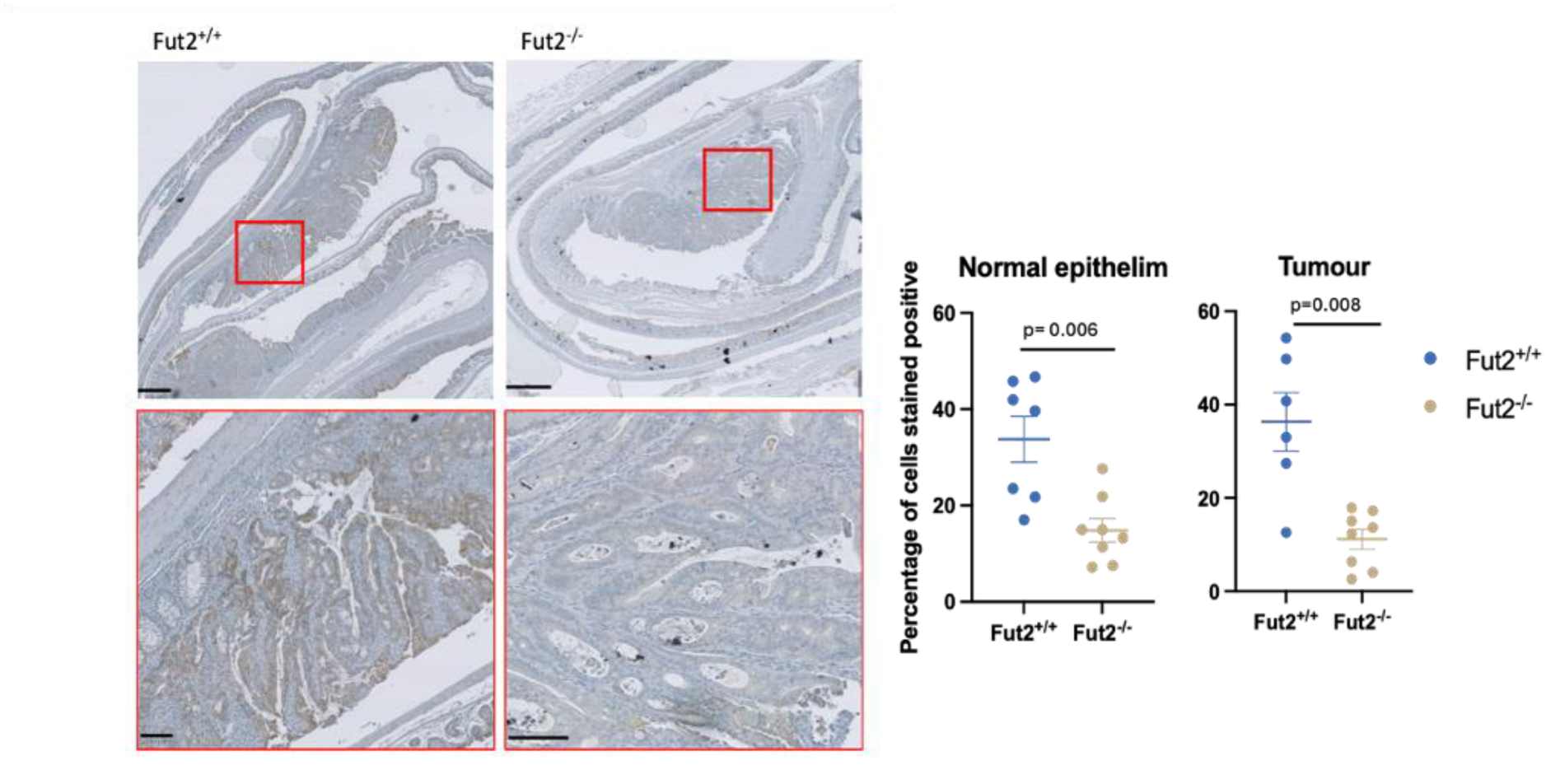
Lower E-cadherin staining in Fut2-/- mice. A Images of Swiss rolls from *Fut2* mice treated with AOM/DSS, stained with Harris Haematoxylin and DAB staining for E-cadherin. Scale bars 500μm. Images highlighted in red show increased magnification, scale bars 100μm, to demonstrate specific epithelial staining. B Comparison of percentage of cells stained positive for E-cadherin in Fut2+/+ (n=7) and Fut2-/- (n=8) mice treated with AOM/DSS (from both Group B and D) in both tumour and normal epithelial tissue. P-value calculated with unpaired T-test with Welch’s correction. Mean and standard error shown.

**Figure 7.**
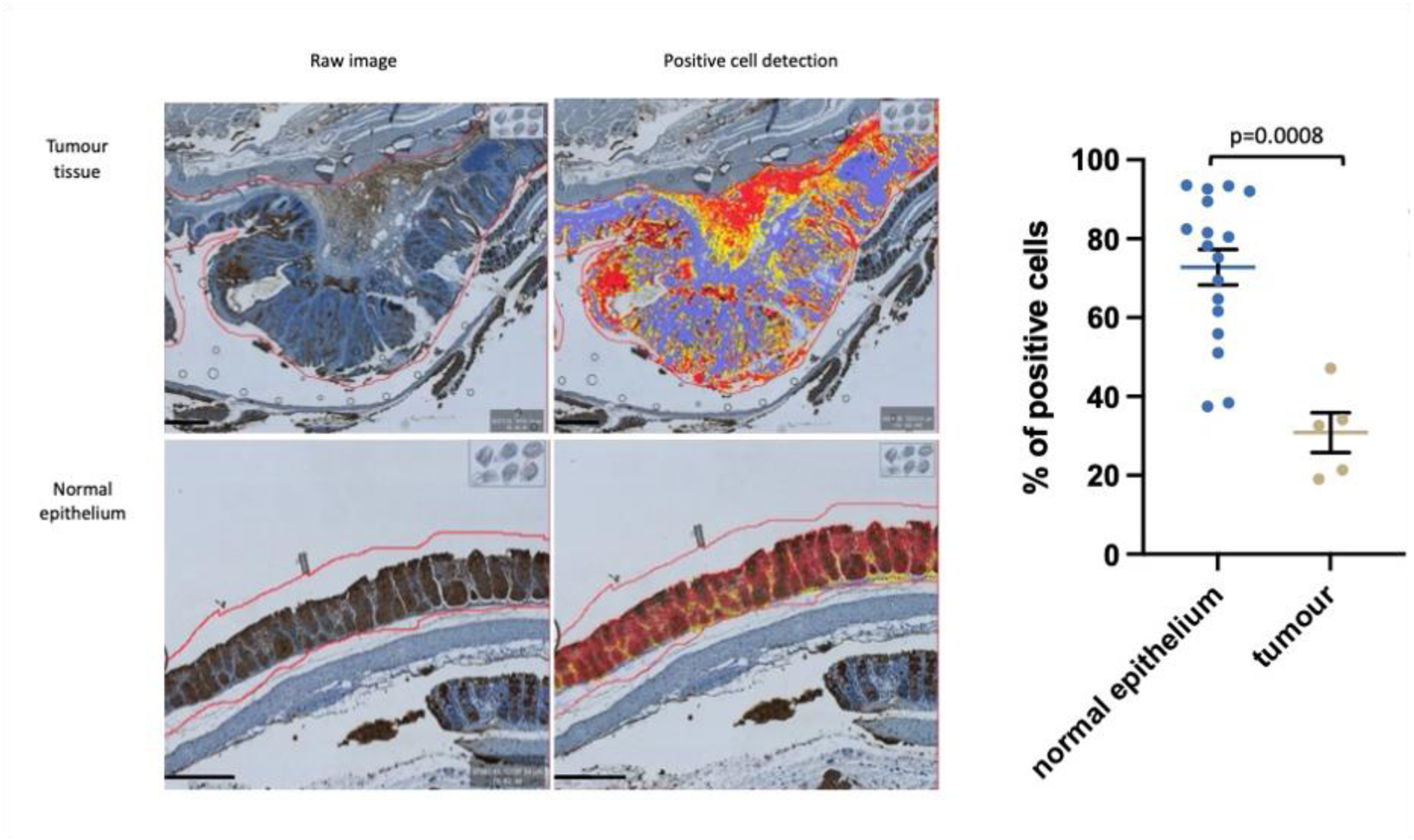
Decreased lectin staining in CRC tissue compared to adjacent normal epithelium in Fut2+/+ mice treated with AOM/DSS. A Representative images of CRC tissue and adjacent normal epithelium stained with UEA-I Lectin (DAB brown-stain), from a *Fut2+/+* mice treated with AOM/DSS. Qu-path “positive cell detection” tool used to analyse the number of cells which stain positively for UEA-I Lectin (highlighted in red). Scale bars shown are 200μm. B Comparison of percentage of cells stained positive for Lectin in normal epithelium and tumour tissues (p=0.0008) P-value calculated with unpaired T-test with Welch’s correction. Mean and standard error of mean shown.

### Loss of *Fut2* decreases survival in an *Apc^min^* murine model

A *Fut2/Apc^min^*murine model was generated in order to interrogate *Fut2* as a CRC- risk locus within the classical CRC pathway. During this breeding process, it was observed that very few *Fut2^-/-^Apc^min/+^* mice were born and surviving beyond 5 days in comparison to *Fut2^-/+^ Apc^min/+^* and *Fut2^+/+^ Apc^min/+^* littermates. Analysis of offspring surviving beyond 5 days revealed a significant underrepresentation of the *Fut2^-/-^Apc^min/+^* mice relative to expected Mendelian ratios (Hardy–Weinberg equilibrium, *p*=0.0135) (Fig. 8).

**Figure 8.**
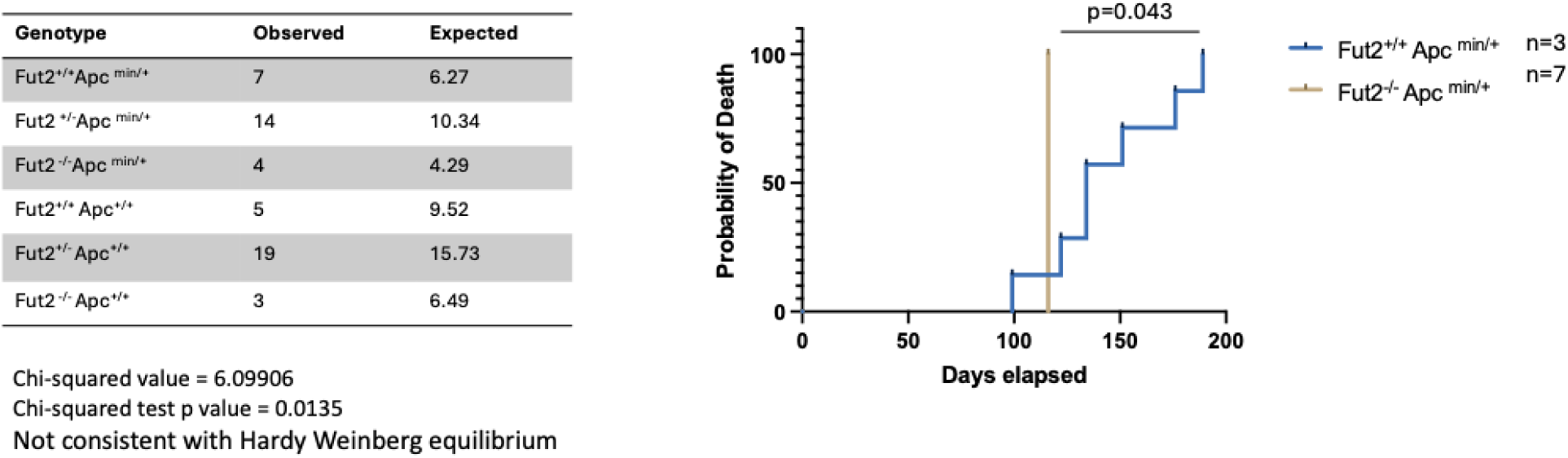
*Fut2-/-Apcmin/+* mice exhibit a greater probability of death. Hardy Weinberg Table depicting observed and expected genotypes for all mice which survived beyond day5 of life. Pearson’s Chi-square test to evaluate deviation from the expected Hary-Weinberg equilibrium in the *Apcmin Fut2* mice. Probability of Death in the *Apcmin Fut2* mouse line with significant differences between the genotypes (p=0.043). Mice were excluded from analysis if the cause of death was unmistakeably due to alternate cause e.g. overgrown teeth, wounds from fighting. P-value calculated using Log rank Mantel Cox Test.

Mice were aged until intestinal clinical phenotypes were observed; pale paws (indicative of anaemia) and hunched posture, significant rectal bleeding, or rectal prolapse. Survival analysis demonstrated that, despite the limited small sample size of *Fut2^-/-^ Apc^min/+^* mice (n=3, all female), this genotype was associated with an increased probability of death compared to Fut2^+/+^Apc^min/+^ mice(n=7) (p=0.043). Mice were excluded from analysis if the cause of death was unmistakeably due to an alternative cause (e.g. overgrown teeth, or injuries resulting from fighting).

When comparing *Fut2^-/+^ Apc^min/+^/Fut2^+/+^ Apc^min/+^* mice with *Fut2^−/−^ Apc^min/+^* mice, there was no significant difference in tumour number, tumour burden, or burden score when males and females were analysed together, indicating no overall genotype effect in the combined cohort. However, when stratified by sex, a statistically significant increase in tumour number and burden was observed in female *Fut2^−/−^ Apc^min/+^* mice compared with female *Fut2^-/+^ Apc^min/+^/Fut2^+/+^ Apc^min/+^*control mice (Fig. 9). This sex-specific effect must be interpreted cautiously, as only three female *Fut2^−/−^ Apc^min/+^* mice were available for analysis, limiting statistical power and increasing susceptibility to sampling variability. Therefore, while the female-only analysis suggests a potential genotype-associated difference, the absence of any effect in the combined cohort indicates that this finding is not robust across the full dataset and requires validation in a larger, sex-balanced cohort.

**Figure 9.**
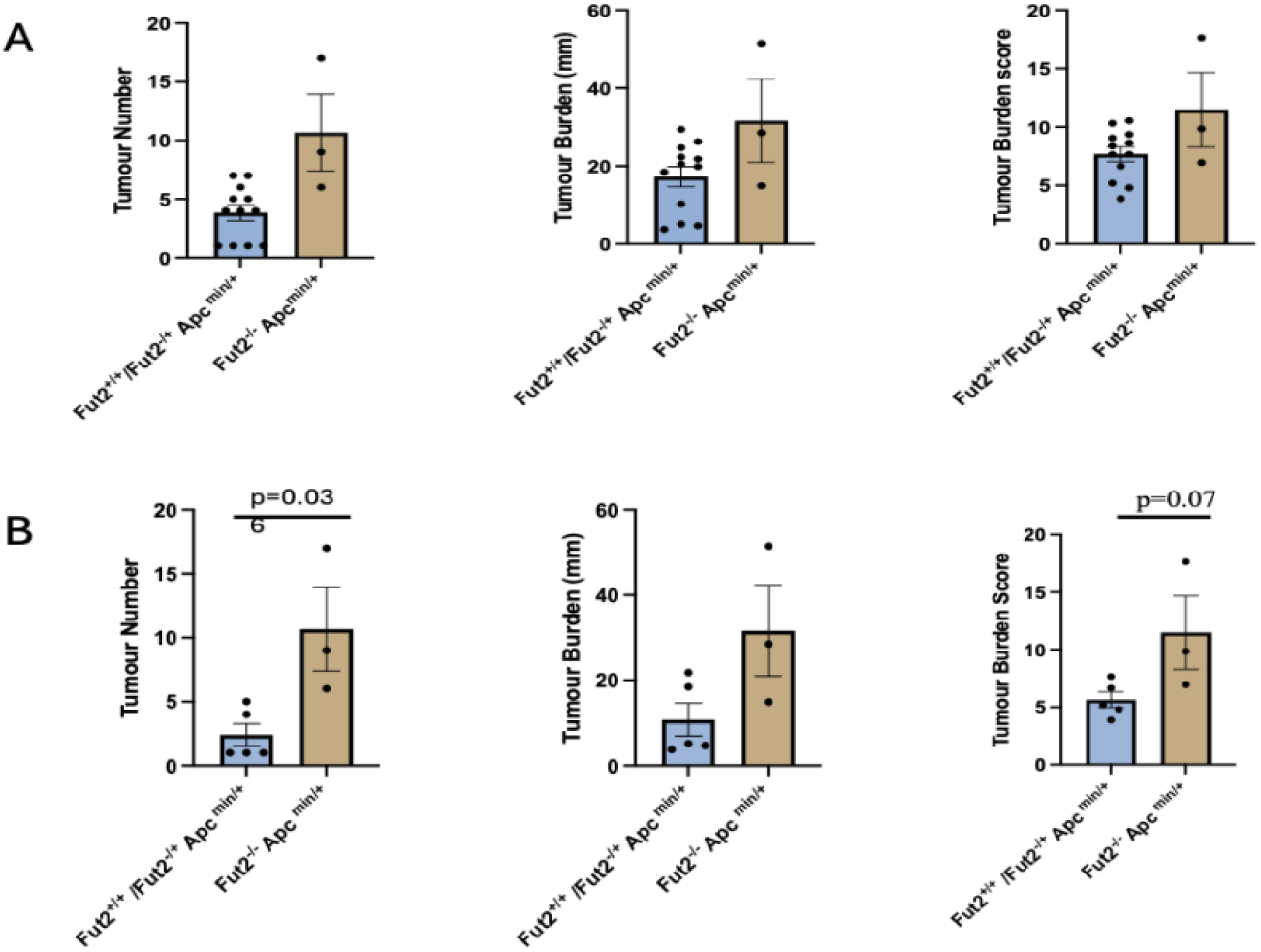
Significant increase in tumour number and burden score in the female *Fut2^-/-^Apc^min/+^*compared to the *Fut2^+/-^/Fut2^+/+^ Apc ^Min/+^*female mice. A. Tumour number, burden (mm) and burden score in *Fut2^+/-^/Fut2^+/+^ Apc ^Min/+^* (n= 5 females, n=7 males) vs. *Fut2^-/-^Apc^min/+^*(n=3 females) of both sex. B Tumour number, burden (mm) and burden score in *Fut2^+/-^/Fut2^+/+^ Apc ^Min/+^* (n=5 females) vs. *Fut2^-/-^Apc^min/+^* (n=3 females) of female only mice.

### The tumour burden is mirrored by B12 supplementation in an inflammatory murine CRC model

A pilot study was conducted to determine the optimal B12 dose for modulating plasma levels in mice. Over four weeks, administration of 100 mg/kg/day resulted in a two-fold increase in plasma B12 concentrations in *Fut2^+/-^* mice (n=4). No phenotypic abnormalities or signs of toxicity were observed at this dose, supporting its use in subsequent experiments.

In the B12 supplementation cohort (Group C; *Fut2^-/-^*n=8, *Fut2^+/+^* n=8), no mice exhibited phenotypic abnormalities or developed tumours spontaneously on macroscopic and microscopic examination. However, microscopic analysis by a colorectal clinical pathologist suggested that three *Fut2^-/-^*mice displayed features suggestive of mononuclear chronic inflammatory cell infiltration within the lamina propria, a finding not observed in *Fut2^+/+^* mice (Fig. 10).

**Figure 10.**
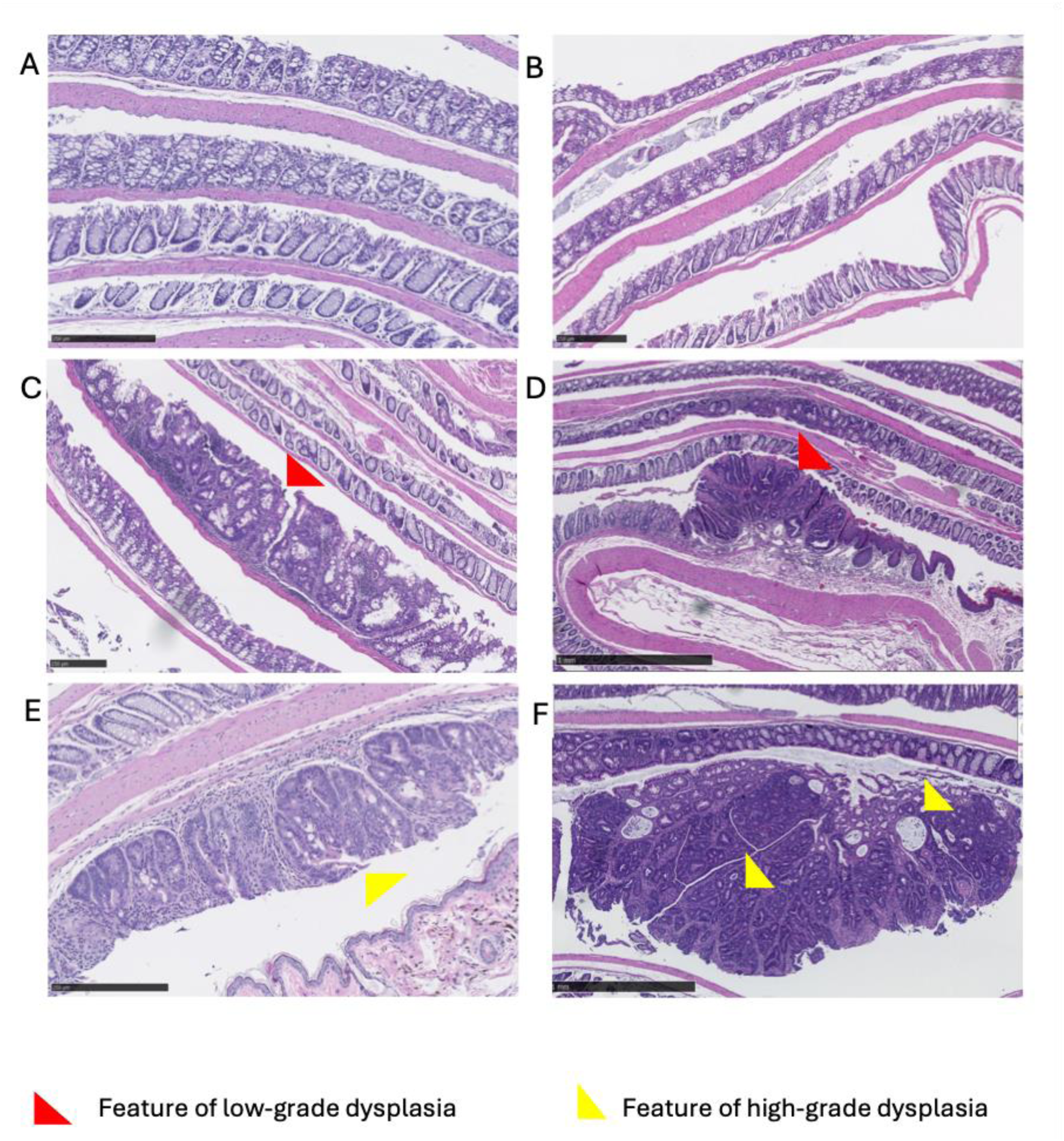
Increased dysplasia seen in AOM/DSS mice treated with Vitamin B12 on histological analysis of H&E-stained Swiss rolls. A *Fut2+/+* B12 only (Group C). The sections show large bowel mucosa with no significant architectural distortion or inflammation. There is no dysplasia. B *Fut2-/-* B12 only (Group C). The colonic mucosa shows no significant architectural distortion. There are possibly focal areas with expansion of mononuclear chronic inflammatory cells within the superficial lamina propria, but this could be spurious due to the plane of section. There is no significant active inflammation and no dysplasia. C *Fut2+/+* AOM/DSS (Group B). There is one small central focus of dysplasia (closer to the distal margin) characterised by closely packed glands, nuclear hyperchromasia and nuclear enlargement. This would be considered as low-grade dysplasia. D *Fut2-/-* AOM/DSS (Group B). There is a tubular adenoma at the squamocolumnar junction. There is some gland crowding but no definite luminal bridging of dysplastic epithelium. The dysplasia is best regarded as being at the upper end of low-grade with no unequivocal high-grade dysplasia seen. E *Fut2+/+* B12/AOM/DSS (Group D). There is one established tubular adenoma just above the squamocolumnar junction. There is increased architectural complexity within the glands with some luminal bridging and fusion and there is focal luminal necrotic debris. The overall features are considered to just amount to high grade dysplasia. F *Fut2-/-* B12/AOM/DSS (Group D). The tubular adenoma exhibits high grade dysplasia characterised by fused, bridged and cribriform glands with luminal necrotic debris, increased nuclear stratification and hyperchromasia with loss of basal polarity. There is some stromal streaming/atypia but no frank desmoplasia. The muscularis mucosae is intact. There is slight mucosal thickening in other areas with increased hyperchromasia within some epithelial cells. Scale bars are to 1mm.

In the B12 supplementation/chemical carcinogen cohort (Group D - B12/AOM/DSS; Fut2^-/-^n=12, *Fut2^+/+^* n=12) mice developed more colonic tumours and had a higher tumour burden score than those treated with AOM/DSS alone. This increased trend was observed in tumour burden score of both genotypes but only reached statistical significant in the *Fut2^+/+^* mice (2.85 (AOM/DSS) vs. 6.48 (B12/AOM/DSS), p=0.028), whilst in the *Fut2^-/-^*mice only a trend was seen (6.38 (AOM/DSS) vs. 8.36 (B12/AOM/DSS0, p=0.148). Indeed, the tumour burden score in the *Fut2^+/+^*mice treated with B12/AOM/DSS was equivalent to the tumour burden score of the *Fut2^-/-^*mice treated with AOM/DSS alone (6.48 (*Fut2^+/+^* B12/AOM/DSS) vs 6.38 (*Fut2^-/-^*AOM/DSS)), consistent across tumour number and burden as well (Fig. 11). Survival probability did not differ between genotypes in the B12/AOM/DSS group, contrasting with decreased survival in *Fut2^-/-^* mice in the AOM/DSS group. Tumour location also varied between the two experimental groups (AOM/DSS and B12/AOM/DSS) with an increased tumour number in the distal section of the colon in the latter.

**Figure 11.**
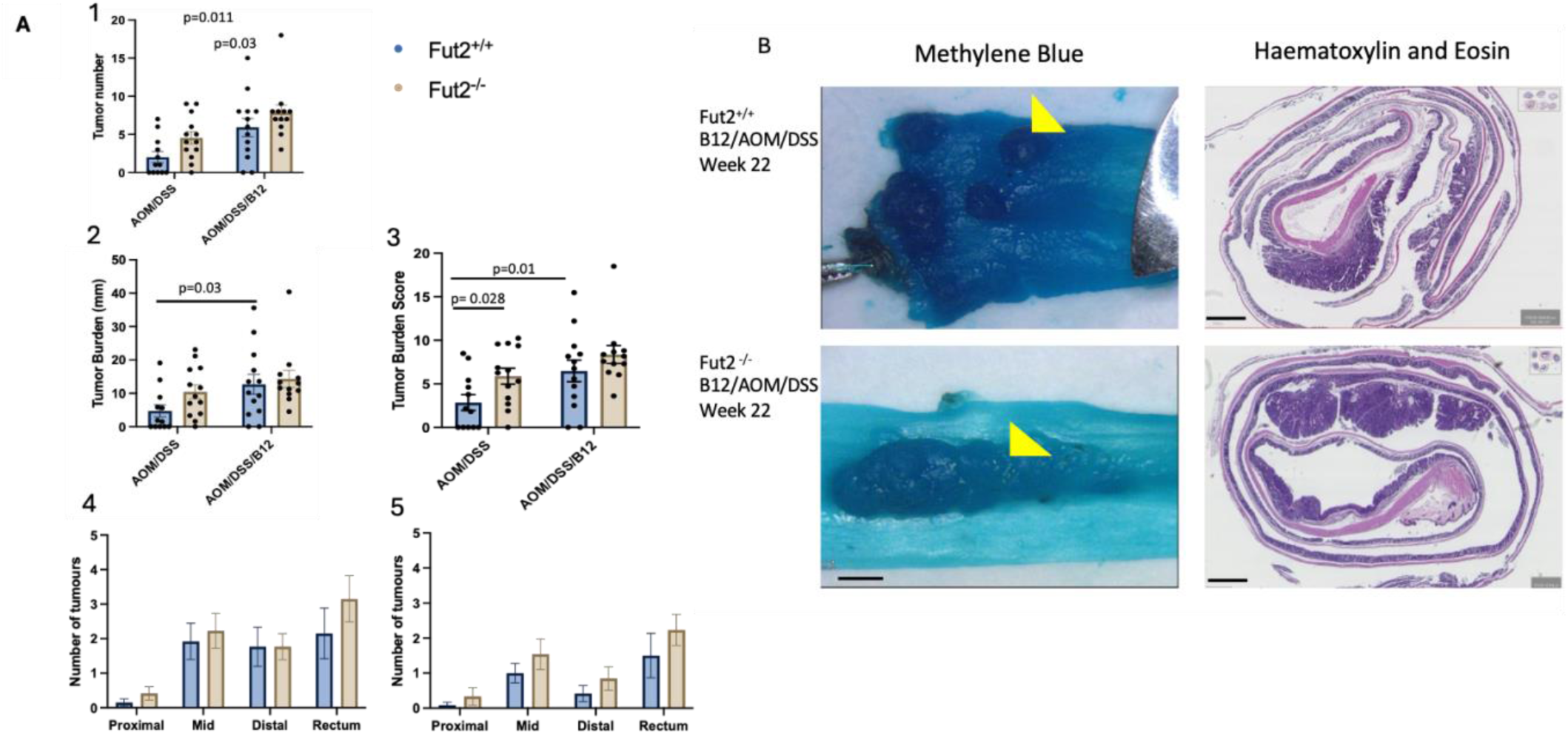
The tumour burden is phenocopied by B12 supplementation in an inflammatory CRC model. **A** Tumour number (1), tumour burden (2), tumour burden score (3) and tumour location (for both B12/AOM/DSS (4) and AOM/DSS (5) with results shown as mean with standard error of mean. P-values calculated with unpaired T-test with Welch’s correction. **B** Sections of colons, stained with methylene blue to highlight the colonic tumours, from Fut2^+/+^ and Fut2^-/-^ mice treated with AOM/DSS and B12 supplementation. H&E sections from the same mice to highlight the cross-sectional view. Yellow arrows indicate tumours.

All intestinal tissues were paraffin-embedded, H&E stained and analysed to confirm that methylene blue–positive areas corresponded to colorectal tumour tissue. A subset of sections were evaluated by a colorectal pathologist, confirming that all tumours were adenomas exhibiting features of either low- or high-grade dysplasia. *Fut2^-/-^* AOM/DSS mice displayed features at the upper limits of low-grade dysplasia whilst *Fut2^+/+^*AOM/DSS mice showed typical low-grade dysplastic changes. In contrast, tumours from the AOM/DSS/B12 group showed features of high-grade dysplasia, irrespective of genotype (Fig 10). Gland crowding was defined as a low-grade feature, whilst high-grade features included luminal bridging, cribriform glands, focal luminal necrotic debris and hyperchromasia with loss of basal polarity. None of the adenomas breached the muscularis mucosae and therefore were not classified as invasive.

### No tumours developed in mice on long-term B12 supplementation

Mice that were treated with 100mg/kg/day of B12 in the drinking water for 1 year (Group E, n=8, male and female) did not develop any tumours. There was no difference in survival, and only one mouse, *Fut2^-/-^* male mouse, had to be culled early do to hunched posture and >20% weight loss.

### No increased tumor burden in *Apc^flox/+^* mice treated with B12

No tumours were observed in control mice by 23 weeks. At the study endpoint, there was no significant difference in tumour number or burden between *Apc^flox/+^*mice receiving B12 supplementation and those given standard drinking water (Fig. 12).

**Figure 12.**
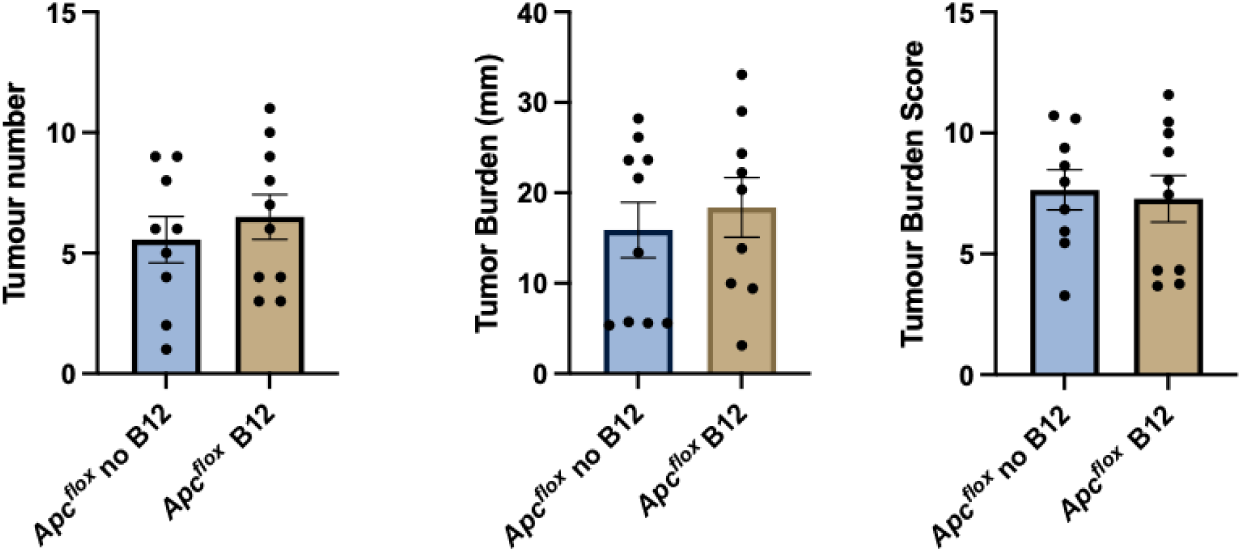
No difference in tumour burden in *Cre+/Apcflox/+* mice treated with or without B12. Tumour number, tumour burden and tumour burden score for with results shown as mean with standard error of mean. P-values calculated with unpaired T-test with Welch’s correction. Figure legend on right refers to all graphs except pilot study.

Survival analysis demonstrated no difference between groups as was the same for clinical scoring at experimental end-point. In the B12-treated *Apc^flox/+^* cohort, two mice were culled prematurely at 20 and 22 weeks due to clinical deterioration (pale paws, hunched posture, and weight loss). In the untreated *Apc^flox/+^* cohort, one mouse was culled at 22 weeks for similar clinical indications.

## Discussion

This body of work validates that the GWAS SNP 19q13.33 (Law et al., 2019), which downregulates the expression of FUT2, is a CRC-risk locus in an *in vivo* model. Variance at 19q13.33 is common, 22-25% of the population are homozygous for the risk allele, even though this locus is only modestly associated with increased CRC-risk (OR=1.07). Therefore, it is not surprising that downregulation of *Fut2* alone was not sufficient to cause spontaneous tumorigenesis in the mice. However, with the addition of inflammatory carcinogens, the tumour burden is significantly higher in the *Fut2^-/-^* mice compared to the *Fut2^+/+^*mice and thus *Fut2* deletion augments the cancer phenotype in an inflammatory neoplastic mouse model.

This is comparable to the results published by Wang et al., albeit the tumour numbers described in that paper are significantly higher (*Fut2^+/+^*=3.5 vs *Fut2^-/-^*=12.2) than the results documented here (*Fut2^+/+^*=2.0 vs *Fut2^-/-^*=4.92).^33^ Furthermore, their survival rate for *Fut2^-/-^* /AOM/DSS mice is significantly lower, with <50% survival by 10 weeks from treatment. This is not surprising given the high dose of DSS the mice were subjected to after AOM injection: 2% DSS for 1 week for a total of 3 cycles in comparison to our study which used 1% DSS for a 1-week course only. Additionally, they counted all tumours, whilst in this study only tumours with a maximum diameter >1mm were considered. This was done to prevent any discrepancies between tumour tissue and lymph nodes, which also stains positive with methylene blue but is commonly <1mm in diameter. In our study by giving a lower DSS dose, there was a greater number of mice surviving until experimental end point and thus comparison of tumour burden between genotypes was more greatly powered, with even animals displaying no tumours retained as true zero values, as the absence of tumour formation is also a meaningful outcome.

These results are in direct contrast to those described by Lieu et al. who found that increased expression of *Fut2* increased the tumour burden.^36^ The model used in this paper is a xenograft model where SW480 cells over-expressing *Fut2* were injected via the hock. This is in essence a very different model, tumours developed at the site of the injection only (the hindlimb) with no colonic tumours and no metastasis. In addition, the study only used female mice (n=6 per genotype), and we found in particular the male mice were more susceptible to tumour formation in our *Fut2-/-* AOM/DSS mice (7.42M vs 4.77F, p= 0.018).

Further validation of 19q13.33 as a CRC-risk locus was then provided by the probability of time to death analysis of the *Fut2/Apc^min^*mice. Although generating sufficient *Fut2^-/-^Apc^min/+^*pups was challenging, the three animals available for analysis still exhibited a significantly increased mortality rate compared to the *Fut2^+/+^ Apc^min/+^* mice. These findings are promising, as they suggest a consistent effect of *Fut2* in a murine CRC model that differs from the inflammatory AOM/DSS model. Despite the differences in model type, the survival data imply a similar directional effect of *Fut2* on disease progression.

In addition to increased tumour burden in the *Fut2^-/-^*mice treated with AOM/DSS, a decrease in E-cadherin staining was also observed. Similar results have previously been noted in mice undergoing intraperitoneal injection with HCT116 cells overexpressing *FUT2* which also showed an increase in E-cadherin staining.^37^ E-cadherin signalling, an essential component of adherens junctions in epithelial cells, undergoes glycosylation modifications and these modifications can influence its stability and function. Reduced E-cadherin expression could reflect a more mesenchymal or dedifferentiated phenotype, which is often observed in the epithelial to mesenchymal transition (EMT) processes, commonly associated with tumour progression. Moreover, E-cadherin signalling is tightly regulated by the Wnt/β-catenin signalling pathway which is known to be dysregulated by lack of O-glycosylation modifications.^38^ Further staining with more EMT markers would need to be carried out to confirm this.

Published literature suggests that increased B12 has been linked to CRC, although to date no *in vivo* models have substantiated this. Furthermore, the interplay between *FUT2* expression, B12 and CRC-risk remains unexplored. This study provides compelling evidence that increased B12 enhances tumorigenicity in a CRC inflammatory murine model. Specifically, the increased tumour burden associated with B12 supplementation was significantly greater in the *Fut2^+/+^* mice than the *Fut2^-/-^* mice and mimicked the tumorigenic phenotype exhibited due to *Fut2* loss. Further, treatment with B12 alone in the *Fut2^-/-^* mice showed a possible increase in the presence of inflammatory cells. This could point to a potential role of B12-mediated increased inflammation in the CRC tumorigenesis.

Conversely, these changes in tumour burden were not mimicked in the *Cre^+^Apc^flox/+^* mice given B12, compared to those treated with normal drinking water, where no differences were seen. This could imply that the effects of B12 are specific to *Fut2* genotype driven changes or are only seen in an inflammatory driven CRC-model.

The molecular mechanism by which B12 may increase tumorigenicity is yet to be established. It has an important role in DNA synthesis, and it may be that increased B12 disrupts this process. Additionally, changes in B12 levels can significantly affect the composition of the intestinal microbiome as it is both synthesized and utilized by the gut microbiome. Interestingly a study which gave mice 10mg/kg/day of B12 in drinking water, used germ-free mice and subsequently orally infected them with *C.rodentium* (a mouse-specific pathogen) during B12 treatment.^39^ The objective of the study was to evaluate the effect of B12 on microbial ecology and results showed that B12 supplementation promoted colonisation with pathogens such as *Parasutterella,* whilst displaying reduced alpha diversity and decreased *Lachnospiraceae* colonies, which are considered symbiotic intestinal bacteria. Mice treated with B12 also showed increased mucosal and epithelial damage compared to control mice. Lurz et al. found, that following DSS induced-colitis (a 5-day course of 2% DSS), mice given B12-deficient chow had a significant decrease in epithelial tissue injury, although no difference in tissue damage between normal and supplemented-chow.^40^ Interestingly they also found >30 genera that were significantly altered in B12-deficient mice, including a decrease in Lachnospiraceae which is the opposite to the study by Forgie et al.

Further evidence to support the notion that B12 increases *FUT2* mediated CRC-risk is through the results of the SMR studies. Importantly these results demonstrated that *FUT2*’s increased CRC-risk is mediated by B12, and this was confirmed in the two independent datasets. The benefit of an SMR study is that it is able to rule out confounding bias by leveraging genetic variants as instrumental variables, thus ensuring that the associations between exposure and outcome are driven by genetic predisposition rather than by external or unmeasured confounders. The difference in effect size between the two studies could be accounted for by differences in the study populations. UKB data is collected from participants UK-wide aged between 40-69, whilst Generation Scotland is participants based only in Scotland aged 18 years and over and GTEx were participants aged 20-70 years old.

While mice provide a valuable *in vivo* model and every effort was made to minimize confounding factors, some unavoidable variables persist, such as cage effects and handling bias, with the occasional change in the technician responsible for the mice. Together, the *in vivo* murine model and the SMR study complement each other, helping to strengthen the overall findings. To conclude, the evidence from these studies demonstrates that *FUT2* deletion augments the cancer phenotype in an established murine model. Supplementation with B12 mimics the additional tumorigenic phenotype due to *FUT2* loss in an inflammatory CRC model, and together with the SMR analysis confirms that a significant proportion of the effect of *FUT2* expression on CRC-risk is mediated by B12.

Results from Forgie et al. and Lurz et al. suggest that this could be associated with B12 modulating the intestinal microbiome. Future studies should focus on interrogating the relationships between B12 and the intestinal microbiome.

## Data Availability

All data produced in the present study are available upon reasonable request to the authors

https://www.gtexportal.org/home

https://shiny.igc.ed.ac.uk/GS_Data_Explorer/

https://www.ukbiobank.ac.uk

## Abbreviations

(AOM): Azoxymethane
(ACME): Average causal mediation effect
(ADE): Average direct effect
(B12): B12
(CRC): Colorectal cancer
(DSS): Dextran sodium sulphate
(eQTL): expression quantitative trait locus
(EMT): epithelial–mesenchymal transition
(GTEx): Genotype-Tissue Expression project
(GWAS): genome-wide association study
(HEIDI): heterogeneity in dependent instruments
(H&E): haematoxylin and eosin
(IHC): immunohistochemistry
(LacZ): β-galactosidase reporter gene
(MR): Mendelian randomisation
(NBF): neutral buffered formalin
(OD): optical density
(PCR): polymerase chain reaction
(RM3): rodent maintenance diet
(SNP): single nucleotide polymorphism
(SMR): summary-data-based Mendelian randomisation
(TCN1): transcobalamin 1
(TCN2): transcobalamin 2
(UKB): UK Biobank
(UEA-I): Ulex europaeus agglutinin I

## Additional Information Ethics

Approval and consent to participate in the clincal arm of the study was granted by the Research Ethics committee and Research and Development teams. REC reference: 23/SS/0062 Protocol number: AC23048 IRAS project ID: 327327

## Data availability

Available at reasonable requests from.

## Acknowledgements

We would like to first acknowledge the excellent management, care and help of Natasha Ackerman and Steven henretty and the rest of the technical staff at the University of Edinburgh BRF/Evans mouse facility. In addition, we would also like to thank the sample processing of the Pathology unit at the Institute of Genetics & Cancer, for their tireless and excellent processing of human and mouse tissues and the Genetics Core at the Wellcome Trust Clinical Research Facility for the shotgun metagenomics seuqencing. We would also like to acknowledge the collection of blood samples from patients by Donna Markie, which was paramount to genotyping ahead of surgery - critical to targeted sample collection. Finally, we would like to offer great appreciation to all the patient donors and their families, without whom this work would not have been possible.

