## Supplementary Tables for "Genetic Variation at 19q13.33 confers colorectal cancer risk through the interaction of mucosal expression of FUT2 and plasma vitamin B12 levels"

| **Name** | **Protocol** | **Sequence (5'-3')** | **Band Size** |
|---|---|---|---|
| *Fut2* Common | JAX Protocol | CCT GCC ATG CTT TCT TTC CTG | Wildtype 154bp<br>Mutant 191bp<br>Heterozygote<br>191bp & 154bp |
| *Fut2* Wild type Reverse | JAX Protocol | ATT CCT TCT CTG ACA GGG TTT GG | |
| *Fut2* Mutant Reverse | JAX Protocol | TGG GTA ACG CCA GGG TTT TC | |

### Supplementary Table 1 List of DNA primers for mice

| **Reagent** | ***Fut2* Reaction Mix (µl)** |
|---|---|
| PCR Mastermix (Thermofisher Scientific) | 5 (1x Conc) |
| dd$H_2O$ | 2.5 |
| DNA | 1 |
| Primers | 0.5 of each (0.5µM) |

### Supplementary Table 2 List of PCR Reaction Mixes used for genotyping mice

| **Temperature (°C)** | **Time** | **Cycle** |
|---|---|---|
| 95 | 5min | |
| 98 | 30s | 20x |
| 65 (-0.5°C/cycle) | 30s | |
| 72 | 45s | |
| 98 | 30s | 20x |
| 55 | 30s | |
| 72 | 45s | |
| 72 | 5min | |
| 10 | hold | |

### Supplementary Table 3 Thermocycler programme for genotyping PCR of *Fut2 mice*

| Clinical Sign | 1 | 2 | 3 |
|---|---|---|---|
| **Rectal Bleeding** | Blood in/on faeces | Visible blood on rectum | Visible blood on fur |
| **Stool Consistency** | Pasty/semi formed/small blood | Diarrhoea not on anus | Diarrhoea adherent to anus |
| **Prolapse** | Mildly apparent | Visible | Symptomatic (Unable to pass faeces) |
| **General Appearance** | Piloerection only | Lethargy, piloerection, intermittently hunched | Motionless, ataxic, sunken eyed, |

**Supplementary Table 4 Clinical scoring system of the mice.** Performed at the time of schedule 1 culling. Mice were culled early if their total score exceeded 4 across all categories, or if they scored 3 or higher in either the Prolapse or General Appearance categories.

| Antibody | Source | Manufacturer | Dilution |
|---|---|---|---|
| Anti E-Cadherin1 | Mouse, monoclonal | BD Transduction Laboratories | 1:500 |
| Anti UEA-I | Rabbit | Sigma Aldrich | 1:500 |

**Supplementary Table 5 Information on Antibodies used for immunohistochemical staining.**

| | Volume (for 1 mouse) | Concentration |
|---|---|---|
| PBS | 25mls | |
| Sodium Deoxycholate | 0.5ml | 5% solution (PBS) |
| NP40 | 50μl | 0.02% solution (PBS) |
| X-gal | 500μl | 1mg/ml |
| Magnesium Chloride | 50μl | 2mM |
| Potassium Ferricyanide | 625μl | 0.07% solution (PBS) |
| Potassium Ferrocyanide | 625μl | 0.09% solution (PBS) |

**Supplementary Table 6 Components of the β-galactosidase staining solution**
